# Seroprevalence of anti-SARS-CoV-2 IgG antibodies in admitted patients at a tertiary referral centre in North India

**DOI:** 10.1101/2022.08.19.22278876

**Authors:** Animesh Ray, Komal Singh, Farha Mehdi, Souvick Chattopadhyay, Ranveer Singh Jadon, Neeraj Nischal, Manish Soneja, Prayas Sethi, Ved Prakash Meena, Anjan Trikha, Gaurav Batra, Naveet Wig

## Abstract

**Background:** Seroprevalence of IgG antibodies against SARS-CoV-2 is an important tool to estimate true burden of infection in a given population. Serosurveys, though being conducted in different parts of India, are not readily published in entirety and often do not report on the different characteristics of the population studied. In this present study, we aimed to serially estimate the seroprevalence of anti-SARS-CoV-2 IgG antibody over 11 months at one of the largest government hospital in India.

**Method:** In this cross-sectional study which was conducted between between 9^th^ June 2020 and 27^th^ April 2021, consecutive patients admitted to medicine wards or intensive care units, who were negative for SARS-CoV-2 by RT-PCR or CBNAAT were included. The 2linic-demographic features of the subjects were recorded in pre-formed questionnaires. Anti-SARS-CoV2 antibody levels targeting recombinant spike receptor-binding domain (RBD) protein of SARS CoV-2 were estimated in serum sample by the ELISA method.

**Results:** A total of 916 patients were recruited over 11 months with mean age(±SD) 39.79±14.9 of years and 55% of population being males. In total 264(28.8%) patients were found to be seropositive. Residency in Delhi and non-smoking status conferred a higher risk for seropositivity. The adjusted odds ratio for seropositivity with regards to no smoking and residence out of Delhi were .31±.09 (Odds ratio ± S.E) and .65 ± .1 (Odds ratio ± S.E) respectively. No other factors like age, socio-economic status, contact history etc showed significant relationship with seropositivity.

**Conclusion:** The seropositivity rate among hospitalized patients was found to increase with time (from 8.45% to 38%) over a period of 9 months. Residence in Delhi and non-smokers had higher risk for seropositivity on multivariate analysis.

## Introduction

After the first case of Corona virus disease 2019 (COVID-19) was detected in India on 30^th^ January 2020, there has been a steady increase of the number of reported cases. (1) India has ranked within the top three countries with the highest case load of diagnosed COVID-19. A meta-analysis had pegged the prevalence of asymptomatic COVID-19 patients between 20-75% in different population. (2) Since asymptomatic COVID-19 cases can be diagnosed early through screening with RT-PCR, a lot of these cases may not diagnosed formally without seroprevalence studies. Seroprevalence studies are thus helpful in gauging the true burden of infection in the community, especially in the background of significant number of asymptomatic cases. Seroprevalence studies for SARS-CoV-2 have been conducted in different settings including community(3), special groups like parturients (4), hemodialysis patients (5), hepatic disease patients (6), blood donors(7), health care workers (8) as well as in hospitals. Very few studies have been however reported from India. As per the first seroprevalence study done in Delhi between 27^th^ June 2020 and 10^th^ July 2021by National Centre for Diseases Control (NCDC) in alliance with Government of National Capital Territory for Delhi, around 22 8% of population of Delhi was seropositive for SARS-CoV-2 (9). The second serosurvey done in the first week of August (10) reported the prevalence at 28.7 %. The third serosurvey (1^st^-15^th^ September 2020) and fourth serosurvey (held in April but truncated due to rapid surge in cases) returned results of 25.1% and 56% respectively.(11) The prime implication of these surveys are that a much larger number of Delhi’s population have been infected with the virus than that revealed by the active case findings by various tests available (RT-PCR, CB-NAAT, rapid antigen test etc). However hospital-based seroprevalence studies have not been commonly done in India. We previously reported the seroprevalence from our centre between June 9^th^ 2020 to August 8^th^ 2020 which was 19.8%(12). Herein, we present the seroprevalence data between the month of June 2020 to April 2021 (before the peak of second COVID-19 wave and subsequent conversion of our hospital to dedicated COVID-19 facility) from our hospitalized patients. We also characterized the clinico-demographic profile of our seropositive patients vis-à-vis seronegative patients.

## Materials and methods

This study was cross-sectional in design and conducted between 9^th^ June 2020 and 27^th^ April 2021 at All India Institute of Medical Sciences, New Delhi among patients admitted to medicine wards and intensive care unit (ICU). Our tertiary care hospital, the largest government hospital in the country, caters to patients from both Delhi and outside (mainly neighbouring states of Haryana, Uttar Pradesh as well as other states in and outside National Capital Region). Consecutive patients (age>14 years) admitted to medicine ward/ICU, whose tests for COVID-19 (RT-PCR or CB-NAAT was negative) was negative at admission, was enrolled in the study. The clinical details of the patients were recorded in a pre-designed questionnaire. The questionnaire, besides capturing demographic details (age, sex, address, state of residence) also enquired about influenza-like symptoms (febrile episodes with one or more of runny nose, sore throat, bodyaches etc where no other alternate diagnosis had been made), comorbidities, addictions (current or ceased within the last three months), diagnosis made for current hospitalization, chest radiograph features etc) After obtaining informed consent, 4 ml of blood was drawn and serum was stored at - 80° C. The test for anti-SARS-Co V-2 antibody was done using ELISA method, developed and validated at Translational Health Science and Technology Institute (THSTI), Faridabad (13), targeting IgG antibody against recombinant spike receptor-binding domain (RBD) protein of SARS-CoV-2. The sensitivity and specificity of the THSTI’s ELISA was higher compared to SARS-CoV-2 IgG ELISAs from Euroimmun and Zydus diagnostics (13). The clinico-demographic details of the patients were captured using preformed case record form filled by trained personnel. A sample size of 800 was estimated by assuming a prevalence of 25%, precision of 3% and alpha-error of 5%.

## Statistical analysis

Categorical variables were calculated as frequency and percentages, continuous variables were represented as mean(± SD). Chi-square test and t test /Wilcoxon ranksum test were used to calculate the statistical differences between categorical variables and continuous variables respectively. A p-value of < .05 was considered to be statistically significant. Categorical variables related to risk factors were recorded and odds ratio (ORs) were calculated using both univariate and mulitivariate analysis by logistic regression to adjust for potential confounders. Multivariate analysis was done using all factors which were either clinically important or having a p value on univariate analysis of atleast .1. In the model similar variables were not analyzed together to avoid collinearity. STATA version 12.1(StataCorp) was used for the statistical analyses required for this study.

## Results

During the study period of 11 months, 916 patients were included in our study. The clinico-demographic details are described in Table 1. In our study population, 55.4% were males and the mean age was 39.79±14.9. Modified Kuppuswamy scale was used to quantify the socioeconomic status of the patients.(14) Majority of the patients belonged to socioeconomic class III(middle) to class V(lower). The vast majority of the patients had one or more comorbidities such as hypertension, diabetes, chronic kidney disease, chronic lung disease, heart disease, past or recently diagnosed tuberculosis. At the time of hospital admission, ∼ 52%(477) were admitted with clinical symptoms pertaining to infective etiology.

**Table 1:** Demographic profile and baseline characteristic of study population

| Variables | Total | IgG Antibody positive (264) | IgG Antibody negative (652) | OR(95% CI) | p Value |
| --- | --- | --- | --- | --- | --- |
| Age | 39.79±14.9 | 40.5±14.89 | 39.48±14.9 | 1.00(0.99, 1.01) |  |
| Male | 506(55.4%) | 146(55.3%) | 360(55.2%) | 1.01(0.76, 1.35) | 0.93 |
| ILI Symptoms | 262(28.6%) | 41(15.5%) | 221(33.9%) | 0.81(0.54, 1.22) | 0.63 |
| Comorbidity | 477(52%) | 125(47.3%) | 352(54%) | 0.86(0.64, 1.15) | 0.095 |
| History of BCG Vaccination | 583(63.6%) | 159(60.2%) | 424(65%) | 0.82(0.61, 1.10) | 0.232 |
| Addiction | 193(21%) | 31(11.7%) | 162(25%) |  | 0.0012 |
| Smoking | 132(14.4%) | 16(6%) | 116(17.8%) | 0.29(0.17, 0.509) | <0.001 |
| Alcohol | 106(12%) | 19(7.2) | 87(13%) | 0.477(0.28, 0.81) | 0.006 |
| Contact | 28(3%) | 5(1.9%) | 23(3.5%) | 0.51(0.19, 1.34) | 0.19 |
| Residence (Delhi) | 513(56%) | 166(63%) | 347(53%) | 1.48(1.11,1.99) | 0.0086 |

**Table 2:**
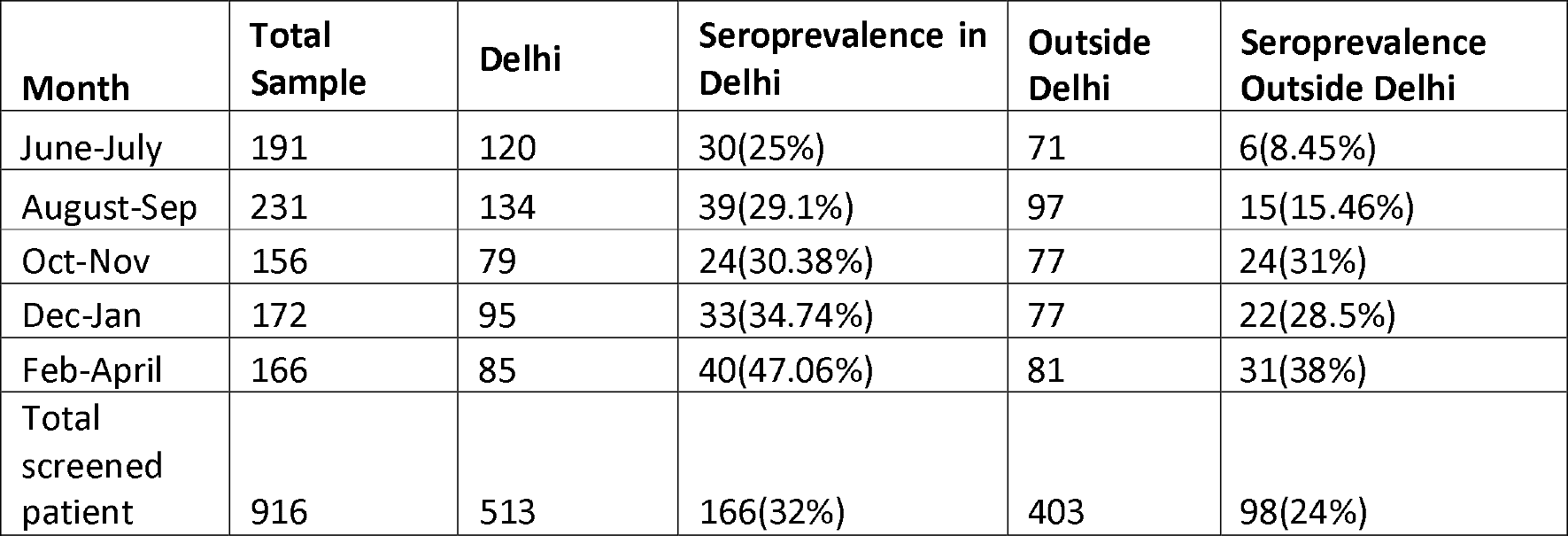
Monthwise distribution of SARS-CoV-2 IgG antibody among patients from Delhi and neighbouring state

**Table 3:** Age wise distribution of SARS-CoV-2 IgG antibody positive case (Age Group 40-49 taken as reference)

| Age Distribution | Antibody (+ve) | Antibody (-ve) | P-value | Relative Risk | 95% CI |
| --- | --- | --- | --- | --- | --- |
| <20 | 19 | 57 | 0.81 | 0.94 | 0.06-1.48 |
| 20-29 | 55 | 150 | 0.89 | 0.97 | 0.70-1.36 |
| 30-39 | 49 | 113 | 0.5 | 1.102 | 0.78-1.54 |
| 40-49 | 48 | 127 |  | 1 |  |
| 50-59 | 62 | 140 | 0.48 | 1.11 | 0.81-1.53 |
| >=60 | 29 | 65 | 0.55 | 1.12 | 0.76-1.65 |

56% of patients were residents of Delhi while the remaining 44% patients were from the states of Uttar Pradesh(23%), Haryana (9.6%), Bihar(5.5%), Rajasthan(0.6%) and others. The month wise seroprevalence data is depicted in Figure 2. The seroprevalence from patients residing both in and out of Delhi rose from June 2020 to April 2021. The differences of seroprevalence between patients from and outside Delhi were significant at all time points. In patients who were seropositive against anti-SARS-CoV-2, no significant differences in history of household contact with COVID-19 cases were noted.

The seroprevalence was highest between 50-59 years and lowest in the <20 years group but the differences in the different age groups were not significant.

On both univariate and multivariate analyses, residence in Delhi and no history of smoking were identified as significant factors determining seropositivity. The adjusted odds ratio for seropositivity with regards to no smoking and residence out of Delhi were 0.31± 0.09 (Odds ratio ± S.E) and 0.65 ± 0.1 (Odds ratio ± S.E) respectively.

25 positive samples on RBD ELISA were retested on Euroimmun ELISA and Zydus Kavach IgG ELISA,out of which 23(92%) was positive on Euroimmun ELISA and 17(68%) was positive on KAVACH ELISA. (THSTI data).

## Discussion

This hospital-based seroprevalence study in the national capital revealed the gradual increase in the percentage of patients demonstrating IgG antibody to SARS-CoV-2. If the population of the study is thought to represent the population of catchment area of the hospital, this study indicates that a substantial proportion of the population of Delhi as well as from the adjoining states to be exposed to the SARS-CoV-2 since the initiation of the pandemic in December 2019. Except for residence inside and outside Delhi, history of no smoking was found to be significantly associated with seropositivity.

The implications of this study are multiple. First, it implies that a significant proportion of population (from which our patient cohort was derived) was seropositive against SARS-CoV-2. With evolution of the pandemic, there was gradual increase in the percentage of seropositive patients. Since, majority of patients have had minor/no symptoms attributable to COVID-19, it signifies that significant proportion of infected patients were asymptomatic and not detected during active case-finding. The result of the recent state-wise sero-survey studies done also lend support to the findings of our study. (11) Next, the seroprevalence data also reflects on the Infection fatality Risk (IFR) of COVID-19. Since, it is not possible to diagnose all cases of COVID-19 infections, the IFR of SARS-CoV-2 has been difficult to determine and has been widely debated (15). The data from this sero survey indicated that the IFR of SARS-CoV-2 in the population of Delhi is ∼0.22 % (12887 deaths reported in Delhi till April 21^st^ 2021)(16), population of Delhi was taken to be 18710922). The crude fatality rate of COVID-19 has been initially thought to be ∼3.4% (17), however the number has been revised several times and appears to shrink further as per the findings of our study. Very low IFR has also been inferred from the seroprevalence studies in other Asian countries including China (8) (with exception of Wuhan), Iran(18) and Israel(19). The national seroprevalence estimate done in India which was initiated in May 2020 had showed a prevalence of infection at 0.73% with an implied IFR of .08% (20). Several reasons for lower IFR in Asian countries including India has been suggested including genetic differences, lower infectious load as well as previous exposure to coronaviruses (21).

The seroprevalence of patients coming from Delhi was significantly higher than that from outside Delhi. This seems to reflect not only the total number of detected cases in Delhi vis-à-vis other states but also the relative percentage of state populated afflicted with COVID-19 infection. The true extent of spread of infection and its surrogate seroprevalence is expected to be proportionate to the percentage of population diagnosed as COVID-19 divided by the number of tests conducted (22).

The relationship between smoking and COVID-19 positivity has been a matter of intense debate. While smoking is known to adversely impact the outcome of COVID-19 patients,(23) current tobacco smoking has been linked to higher risk of asymptomatic (with respect to symptomatic infections)(24)(25)(26) and COVID-19 infections (27) The possible reasons for low seropositivity among smokers may be due to action on nicotinic acetylcholine receptors(nAChR) reducing potential sites for viral entry in the pulmonary alveolar epithelium and through inhibition of pro-inflammatory cytokines.(25)(28) Alternative explanation include more episodes of upper respiratory tract infections in smokers leading to cross-protection afforded by infecting viruses including common cold coronaviruses which have significant genetic similarity to SARS-CoV-2.(29)(24)

Our study were limited by its single centre setting as well as scope for misclassification due to recall bias in the study participants. Also some potential factors like those influencing mixing in society, quantum of smoking and alcohol exposure and other predisposing factors for respiratory infections were not included in our study.

To conclude, in our serosurvey of hospitalized patients, the seroprevalence was found to be gradually increasing with time. Residence in Delhi and non-smoking status were the two important factors predicting seropositivity in our study. Future studies should focus on confirming the findings of this study as well as determine the probable rise in seroprevalence after the occurrence of second COVID-19 wave.

## Data Availability

The data may be made available on request to the authors

---

**FIGURE 1.**
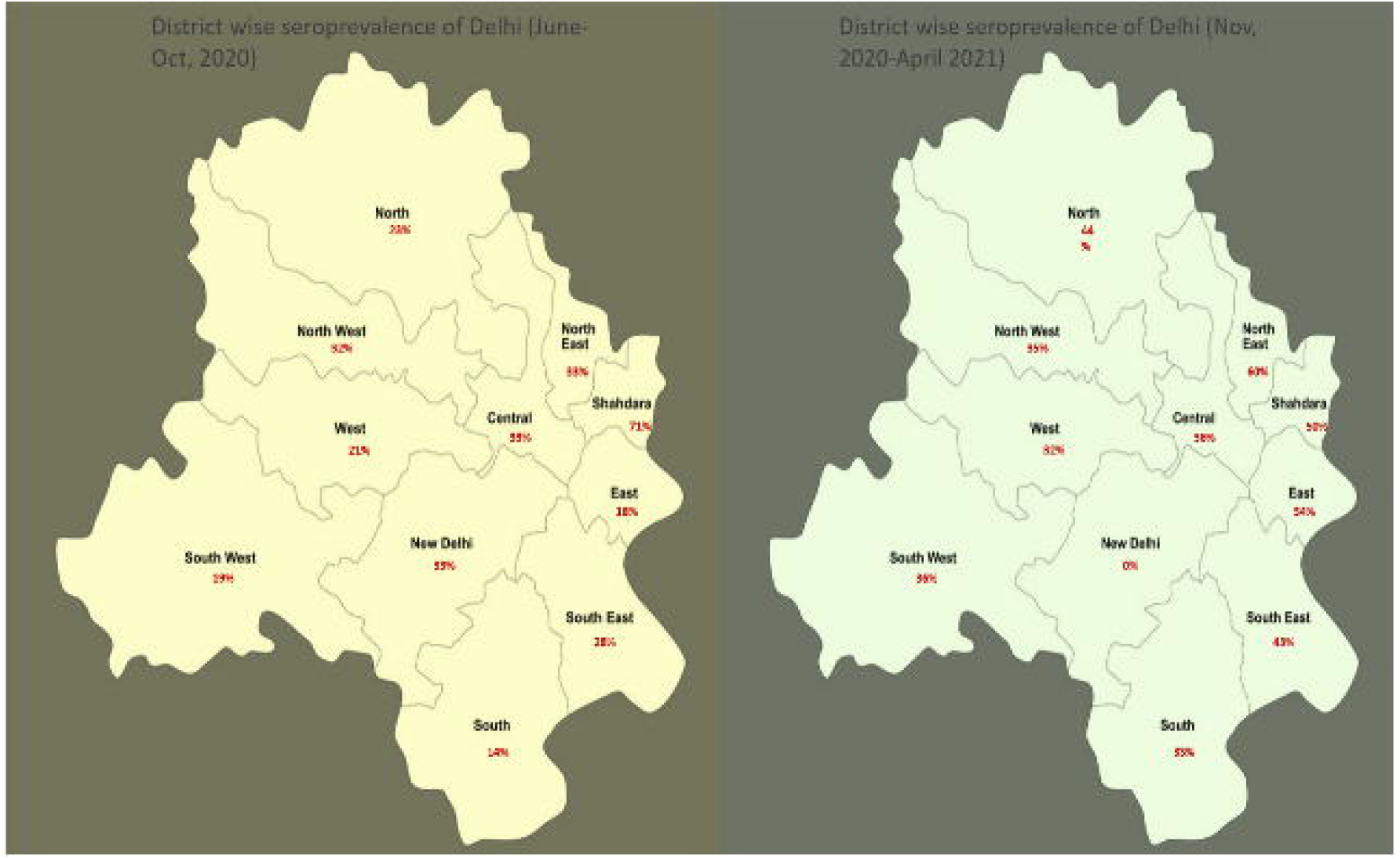

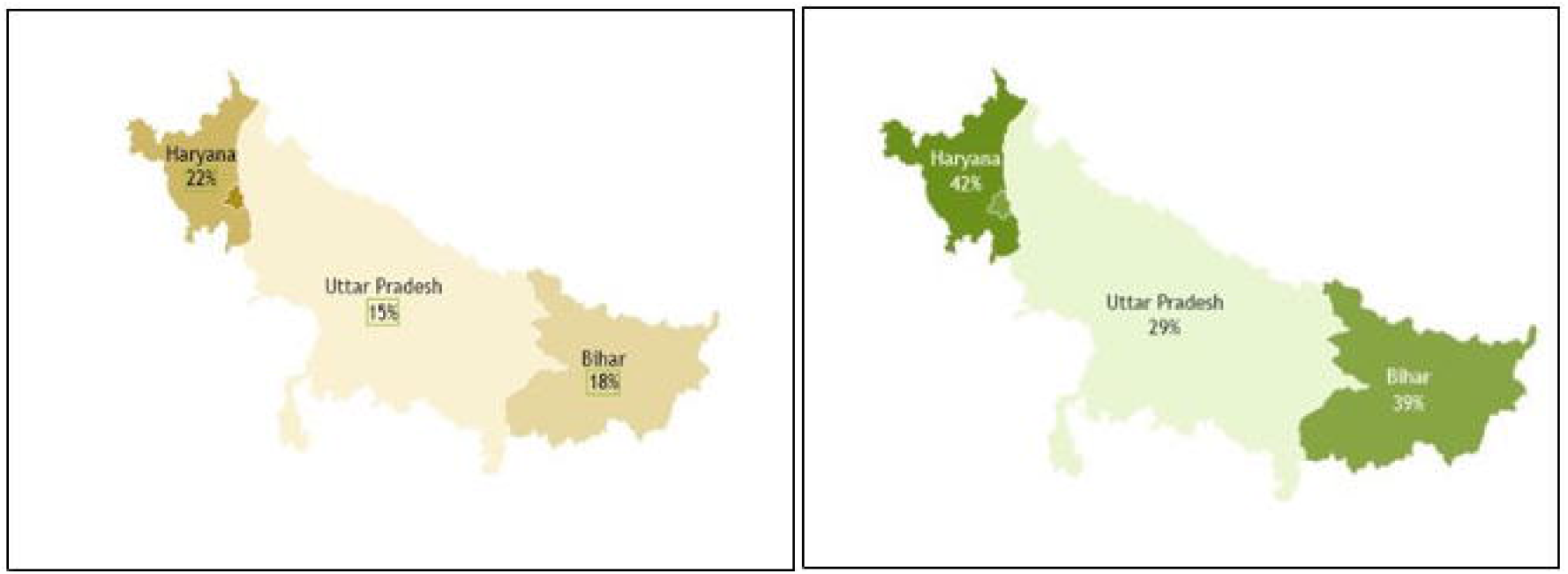

## Bibliography

1. Andrews MA, Areekal B, Rajesh KR, Krishnan J, Suryakala R, Krishnan B, et al. First confirmed case of COVID-19 infection in India: A case report. Indian J Med Res. 2020 May;151(5):490–2.

2. Yanes-Lane M, Winters N, Fregonese F, Bastos M, Perlman-Arrow S, Campbell JR, et al. Proportion of asymptomatic infection among COVID-19 positive persons and their transmission potential: A systematic review and meta-analysis. PloS One. 2020;15(11):e0241536.

3. Pollán M, Pérez-Gómez B, Pastor-Barriuso R, Oteo J, Hernán MA, Pérez-Olmeda M, et al. Prevalence of SARS-CoV-2 in Spain (ENE-COVID): a nationwide, population-based seroepidemiological study. Lancet Lond Engl. 2020 Jul 3;

4. Flannery DD, Gouma S, Dhudasia MB, Mukhopadhyay S, Pfeifer MR, Woodford EC, et al. SARS-CoV-2 Seroprevalence Among Parturient Women. MedRxiv Prepr Serv Health Sci. 2020 Jul 10;

5. Clarke C, Prendecki M, Dhutia A, Ali MA, Sajjad H, Shivakumar O, et al. High Prevalence of Asymptomatic COVID-19 Infection in Hemodialysis Patients Detected Using Serologic Screening. J Am Soc Nephrol JASN. 2020 Jul 30;

6. Suda G, Ogawa K, Kimura M, Maehara O, Kitagataya T, Ohara M, et al. Time-dependent changes in the seroprevalence of COVID-19 in asymptomatic liver disease outpatients in an area in Japan undergoing a second wave of COVID-19. Hepatol Res Off J Jpn Soc Hepatol. 2020 Jul 30;

7. Ng D, Goldgof G, Shy B, Levine A, Balcerek J, Bapat SP, et al. SARS-CoV-2 seroprevalence and neutralizing activity in donor and patient blood from the San Francisco Bay Area. MedRxiv Prepr Serv Health Sci. 2020 May 25;

8. Xu X, Sun J, Nie S, Li H, Kong Y, Liang M, et al. Seroprevalence of immunoglobulin M and G antibodies against SARS-CoV-2 in China. Nat Med. 2020;26(8):1193–5.

9. Sero-prevalence study conducted by National Center for Disease Control NCDC, MoHFW, in Delhi, June 2020 [Internet]. [cited 2020 Aug 17]. Available from: pib.gov.in/Pressreleaseshare.aspx?PRID=1640137

10. Second sero survey in Delhi reveals more people have developed Covid-19 antibodies [Internet]. Hindustan Times. 2020 [cited 2021 Oct 3]. Available from: https://www.hindustantimes.com/delhi-news/second-sero-survey-in-delhi-reveals-more-people-have-developed-covid-19-antibodies/story-Ajkxid6iz9OMi3RlQjo7rN.html

11. Cut short by fourth wave, Delhi’s latest sero study offers few clues [Internet]. Hindustan Times. 2021 [cited 2021 Oct 3]. Available from: https://www.hindustantimes.com/cities/delhi-news/cut-short-by-fourth-wave-delhi-s-latest-sero-study-offers-few-clues-101624486972183.html

12. Seroprevalence of anti-SARS-CoV-2 IgG antibody in hospitalized patients in a tertiary referral center in North India | medRxiv [Internet]. [cited 2021 Sep 26]. Available from: https://www.medrxiv.org/content/10.1101/2020.08.22.20179937v1

13. Frontiers | Development of a Fast SARS-CoV-2 IgG ELISA, Based on Receptor-Binding Domain, and Its Comparative Evaluation Using Temporally Segregated Samples From RT-PCR Positive Individuals | Microbiology [Internet]. [cited 2021 Sep 26]. Available from: https://www.frontiersin.org/articles/10.3389/fmicb.2020.618097/full

14. Saleem SM, Jan SS. Modified Kuppuswamy socioeconomic scale updated for the year 2021. Indian J Forensic Community Med. 2021 Apr 28;8(1):1–3.

15. Faust JS, Rio C del. Assessment of Deaths From COVID-19 and From Seasonal Influenza. JAMA Intern Med. 2020 Aug 1;180(8):1045–6.

16. Number of Cases [Internet]. PRS Legislative Research. [cited 2021 Oct 3]. Available from: https://prsindia.org/covid-19/cases

17. WHO Director-General’s opening remarks at the media briefing on COVID-19 - 3 March 2020 [Internet]. [cited 2020 Aug 9]. Available from: https://www.who.int/dg/speeches/detail/who-director-general-s-opening-remarks-at-the-media-briefing-on-covid-193-march-2020

18. Shakiba M, Hashemi Nazari SS, Mehrabian F, Rezvani SM, Ghasempour Z, Heidarzadeh A. Seroprevalence of COVID-19 virus infection in Guilan province, Iran [Internet]. Infectious Diseases (except HIV/AIDS); 2020 May [cited 2020 Aug 20]. Available from: http://medrxiv.org/lookup/doi/10.1101/2020.04.26.20079244

19. Time course of Case fatality rate (case-fatality ratio) for COVID-19 in Israel [Internet]. [cited 2021 Sep 26]. Available from: https://www.science.co.il/medical/coronavirus/Case-fatality-rate.php

20. India not in community transmission stage, states cannot lower guard, says ICMR [Internet]. The Indian Express. 2020 [cited 2020 Aug 9]. Available from: https://indianexpress.com/article/india/india-not-in-community-transmission-stage-yet-health-ministry-6454183/

21. Ioannidis J. The infection fatality rate of COVID-19 inferred from seroprevalence data [Internet]. Infectious Diseases (except HIV/AIDS); 2020 May [cited 2020 Aug 9]. Available from: http://medrxiv.org/lookup/doi/10.1101/2020.05.13.20101253

22. Shams SA, Haleem A, Javaid M. Analyzing COVID-19 pandemic for unequal distribution of tests, identified cases, deaths, and fatality rates in the top 18 countries. Diabetes Metab Syndr. 2020;14(5):953–61.

23. Patanavanich R, Glantz SA. Smoking Is Associated With COVID-19 Progression: A Meta-analysis. Nicotine Tob Res Off J Soc Res Nicotine Tob. 2020 Aug 24;22(9):1653–6.

24. Saurabh S, Verma MK, Gautam V, Kumar N, Jain V, Goel AD, et al. Tobacco, alcohol use and other risk factors for developing symptomatic COVID-19 vs asymptomatic SARS-CoV-2 infection: a case-control study from western Rajasthan, India. Trans R Soc Trop Med Hyg. 2021 Jul 1;115(7):820–31.

25. Farsalinos K, Barbouni A, Poulas K, Polosa R, Caponnetto P, Niaura R. Current smoking, former smoking, and adverse outcome among hospitalized COVID-19 patients: a systematic review and meta-analysis. Ther Adv Chronic Dis. 2020;11:2040622320935765.

26. Grundy EJ, Suddek T, Filippidis FT, Majeed A, Coronini-Cronberg S. Smoking, SARS-CoV-2 and COVID-19: A review of reviews considering implications for public health policy and practice. Tob Induc Dis. 2020;18:58.

27. Paleiron N, Mayet A, Marbac V, Perisse A, Barazzutti H, Brocq F-X, et al. Impact of Tobacco Smoking on the Risk of COVID-19: A Large Scale Retrospective Cohort Study. Nicotine Tob Res Off J Soc Res Nicotine Tob. 2021 Aug 4;23(8):1398–404.

28. Changeux J-P, Amoura Z, Rey FA, Miyara M. A nicotinic hypothesis for Covid-19 with preventive and therapeutic implications. C R Biol. 2020 Jun 5;343(1):33–9.

29. Ma Z, Li P, Ji Y, Ikram A, Pan Q. Cross-reactivity towards SARS-CoV-2: the potential role of low-pathogenic human coronaviruses. Lancet Microbe. 2020 Aug;1(4):e151.

